# *PALM3* and hearing loss: a potential dual diagnosis interfering with novel gene discovery

**DOI:** 10.64898/2026.04.20.26351093

**Authors:** Paria Najarzadeh Torbati, Lukas Hallbrucker, Michaela A.H. Hofrichter, Daniel Owrang, Jonas Setzke, Manfred W. Kilimann, Anahid Hemmatpour, Mohsen Rajati, Ehsan Ghayoor Karimiani, Thomas Haaf, Christian Vogl, Barbara Vona

## Abstract

Hereditary hearing loss is highly genetically heterogeneous, with emerging overlap between genes implicated in early-onset and age-related hearing loss. We report a consanguineous family with autosomal recessive, non-syndromic hearing loss in which the proband harbors a homozygous splice-site variant in *PALM3* (NM_001145028.2:c.314+1G>A) and a homozygous missense variant in *OTOA*. A minigene assay for the *PALM3* variant demonstrated aberrant splicing with exon skipping, resulting in a frameshift and a large inframe deletion, both consistent with loss of function and impacting all known transcripts. While the organ of Corti from 12-month-old heterozygous *Palm3* mice showed preserved overall architecture, published *Palm3* knockout mice exhibit auditory dysfunction, supporting an auditory phenotype with loss of function. Although a dual molecular diagnosis cannot be excluded, the combined genetic, functional, and comparative data support *PALM3* as a strong candidate gene for autosomal recessive hearing loss.

## INTRODUCTION

Cochlear hair cells convert mechanical sound waves into electrical signals. This process relies on ribbon synapses, electron-dense structures that tether synaptic vesicles to enable sustained neurotransmission (Smith and Sjöstrand, 1961). In mammals, two functionally distinct hair cell types cooperate in auditory processing: inner hair cells (IHCs), which transmit acoustic signals via ribbon synapses, and outer hair cells (OHCs), which amplify cochlear motion through voltage-dependent electromotility (Halim et al., 2025). Each IHC forms monosynaptic connections with multiple auditory nerve fibers, and synaptic heterogeneity contributes to diverse firing properties (Liberman, 1980).

Hereditary hearing loss is highly genetically heterogeneous. More than 155 genes are associated with early-onset non-syndromic hearing loss, while genome-wide association studies have identified approximately 80 candidate genes for age-related hearing loss (ARHL), despite many causative genes still pending discovery (Cornejo-Sanchez et al., 2025). Although ARHL genetics has not been used for routine diagnostics yet, previous studies on twins and families indicated that genetic factors play a key role, for approximately 50-70%, in ARHL (Kvestad et al., 2012; Duran et al., 2025). These genetic regions highlight biological pathways that overlap with Mendelian deafness genes and suggest further in-depth studies are needed (Wells et al., 2019).

Paralemmins are a group of membrane protein families, PALM1-3–3, and Palmdelphin, which contribute to membrane shaping and cytoskeletal interactions (Kutzleb et al., 1998; Hultqvist et al., 2012). A recent study demonstrated that *Palm3* is expressed in both IHCs and OHCs and concentrates at their lateral membranes. *Palm3*-knockout (KO) mice exhibit early-onset, progressive hearing loss due to shortening of OHCs, membrane disorganization, and cytoskeletal disruption. Adeno-associated virus-mediated *Palm3* delivery partially restored structure and function (Halim et al., 2025).

Here, we report a family with autosomal recessive (AR) hearing loss harboring the first known biallelic loss-of-function (LOF) variant in *PALM3*. The affected individual also segregates a homozygous missense variant in the established deafness gene, *OTOA* (DFNB22), suggesting a potential dual diagnosis consistent with the extreme heterogeneity of hereditary hearing loss. Functional analysis demonstrates *PALM3* exon skipping, and phenotypic concordance with *Palm3*-KO mice supports a LOF mechanism. These findings implicate *PALM3* as a candidate human deafness gene essential for hair cell membrane resilience and cochlear function.

## METHODS

### Patient recruitment and clinical assessment

We evaluated an extended consanguineous family comprising two branches with suspected different causes of hearing impairment. This family was included in a large ethnically diverse rare disease study consisting of approximately 800 probands with the sole inclusion criteria being hereditary hearing impairment. This study was approved by the Medical Faculty of the University of Würzburg, Germany (approval numbers 131/21, 226/18 and 46/15). Blood samples were collected after obtaining informed consent from patients or their parents. Written informed consent from the parents or legal guardians of the patients/participants was obtained for the publication of their data.

Demographic, otolaryngologic, audiological, and relevant medical data were ascertained from the medical records of the proband (Figure 1A, arrow) and spouse. Affected individuals underwent a complete otologic evaluation. Routine pure-tone audiometry was performed according to current standards and measured hearing thresholds at 0.25, 0.5, 1, 2, 4, and 8 kHz. Air- and bone-conduction thresholds were measured and severity of hearing loss was determined by averaging pure-tone thresholds over 0.5, 1, 2 and 4 kHz (pure-tone average, PTA_0.5-4K_).

**Figure 1.**
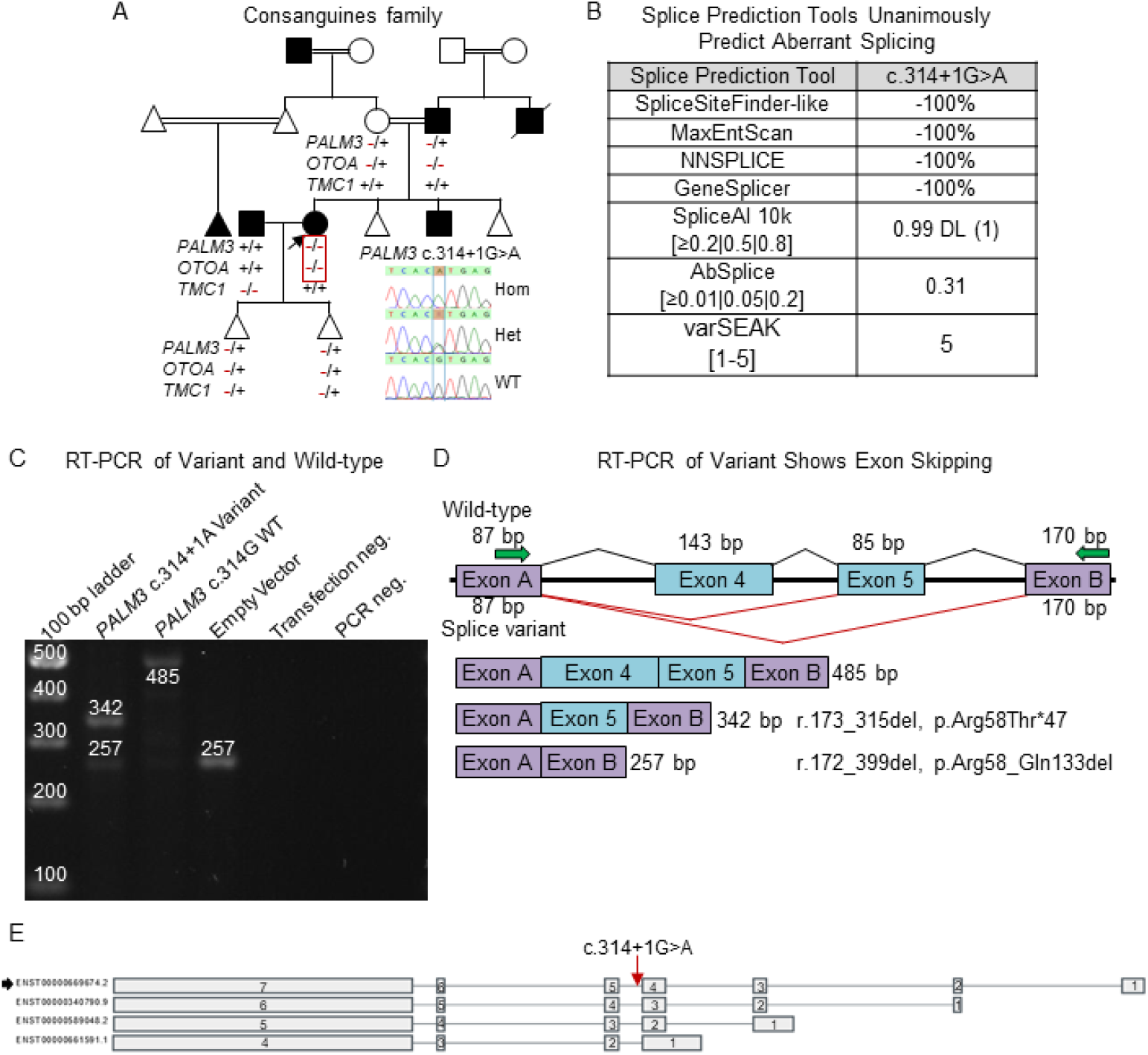
Pedigree, *in silico* splice predictions, *in vitro* splice assay and transcript analysis. A. The pedigree of the consanguineous family showing segregation of the *PALM3* c.314+1G>A, p.?, *OTOA* c.1939G>C, p.(Gly647Arg), and *TMC1* c.1535G>A, p.(Arg512Gln) variants. The Sanger sequencing inset shows the electropherograms of the *PALM3* c.314+1G>A, p.? variant. B. Results of the *in silico* splice prediction tools with unanimous aberrant splicing predicted. C. RT-PCR of the *in vitro* splice assay testing the *PALM3* c.314+1G>A variant. The variant (c.314+1A) shows two bands corresponding to skipping of exon 4 and exons 4-5, while the wild-type (c.314+1G), empty vector, transfection and PCR controls performed as expected. D. A schematic of the results of the *in vitro* splice assay. The upper panel shows the wild-type splicing result containing the expected exons 4-5 joined with black lines. The bottom panel shows the skipped exon splicing results. Skipped exon 4 and 4-5 are joined with red lines. The illustration below shows the splicing results from the agarose gel showing the RT-PCR in panel C with the amplicon sizes shown. The aberrant splicing results are shown with their r. and p. positions. E. The aberrant splice effect would impact all four human *PALM3* transcripts.

### Exome sequencing, bioinformatics and variant classification

The genomic DNAs of the proband and their spouse were exome sequenced. Following extraction of DNAs from whole blood by standard protocols, proband DNA samples were subjected to exome capture using the Nextera DNA Exome (Illumina, Inc., San Diego, CA, USA) according to manufacturer’s protocols and paired-end sequenced on an Illumina sequencer. The exome data were analyzed and processed as previously described (Redfield et al., 2024). Copy number variation analysis was performed. A focus on hearing loss-associated genes was first prioritized in exome analyses and later included exome-wide investigations.

The generated sequences were demultiplexed and mapped to the human genome reference (GRCh38) with Burrows Wheeler Aligner for subsequent variant calling. Bioinformatics filtering strategy focused on exonic and donor/acceptor splicing variants. Alternative alleles present at >20% and a minor allele frequency <0.01 were assessed using gnomAD (Chen et al., 2024), TopMed (Taliun et al., 2021) and the All of Us research program (The All of Us Research Program Genomics Investigators et al., 2024). In silico pathogenicity predictions applied SIFT (Ng and Henikoff, 2003), PolyPhen-2 (Adzhubei et al., 2010), FATHMM (Shihab et al., 2013), MutationTaster (Steinhaus et al., 2021), REVEL (Ioannidis et al., 2016), and CADD (Schubach et al., 2024). Computational assessment of splicing effects used SpliceSiteFinder-like, MaxEntScan, NNSplice, and GeneSplicer embedded in Alamut Visual Plus v1.6.1 (Sophia Genetics, Bidart, France), as well as varSEAK (JSI Medical Systems), SpliceAI Visual and AbSplice embedded in SpliceAI Visual (De Sainte Agathe et al., 2023).

Pathogenicity of identified variants in known hearing loss-associated genes was evaluated according to the hearing loss-adapted American College of Medical Genetics/Association for Molecular Pathology (ACMG/AMP) guidelines (Oza et al., 2018). Variants were analyzed in ClinVar and the Deafness Variation Database (DVD) (Azaiez et al., 2018) to assess previous variant classifications. Variants were subjected to segregation analysis by Sanger sequencing with the DNA samples from available family members. Primers are shown in Supplementary Table 1. Visualization of truncated variants and variant nomenclature validation was performed using Mutalyzer (Lefter et al., 2021).

### *In vitro* splice assay

RNA studies of variants were conducted following established protocols with some modifications (Tompson and Young, 2017; Rad et al., 2021). In brief, the minigene construct comprised an 864 bp region containing exons 4 and 5 with flanking intronic sequence. This region was amplified from the proband and a healthy control using primers containing specific restriction sites. All primers are shown in Supplementary Table 1. The PCR fragments were ligated between exons A and B of the linearized pSPL3-vector following digestion with restriction enzymes. The recombinant vectors were transformed into DH5_α_ competent cells (NEB 5-alpha, New England Biolabs, Frankfurt, Germany), plated and incubated overnight. After conducting colony PCR with pSPL3 Primer SD6 F and the cloning target-specific reverse primer, the results were analyzed. The sequences of both the wild-type and mutant-containing vectors were confirmed through Sanger sequencing. Subsequently, these vectors were transfected into HEK293T cells (ATCC, Manassas, VA, USA). Transfection was achieved by introducing 2 µg of the respective pSPL3 vectors using 6 µL of FuGENE 6 Transfection Reagent (Promega, Walldorf, Germany). The empty vector control was also included in the experiment as control. The cells were then harvested 24 hours post-transfection, and total RNA was isolated using the miRNeasy Mini Kit (Qiagen, Hilden, Germany). cDNA was synthesized using the High Capacity cDNA Reverse Transcription Kit (Applied Biosystems, Waltham, MA, USA) following the manufacturer’s protocols. cDNA was PCR amplified using vector-exon specific primers. The amplified fragments were visualized on a 1% agarose gel.

### Immunohistological Analysis of Cochlear Hair Cells in *Palm3*^*+/*−^ Mice

*Palm3* heterozygous mice (12 months old) and wild-type mice (11 months old) were euthanized, and dissected cochleae were fixed for 1 h at 4 °C in 4% formaldehyde in PBS. Cochleae were decalcified in 0.3 M EDTA for 3 days with repeated solution changes. Apical– medial turns of the organ of Corti were then dissected for subsequent immunohistological staining (Halim et al., 2025).

Tissues were permeabilized for 30 min in permeabilization buffer (0.5% Triton X-100 in PBS) and blocked for 1 h in blocking buffer (10% goat serum in permeabilization buffer). Samples were incubated overnight at 4 °C with anti-Myosin7A antibody (MYO7A 138-1, DSHB; 1:500 dilution), followed by a 3-h incubation at room temperature with goat anti-mouse IgG1 Alexa Fluor 488 (A-21121, Invitrogen; 1:500 dilution) and TO-PRO-3 iodide (T3605, Invitrogen; 1:1000 dilution). Tissues were mounted on glass slides using a Mowiol-based mounting medium prior to imaging.

Hair cells were counted in a 300-µm-long region of interest located 1600 µm from the apex of the organ of Corti, measured along the inner hair cell row. Quantification was performed using FIJI/ImageJ.

## RESULTS

### Case report

The adult proband with bilateral, sensorineural hearing loss was born to consanguineous parents. No vestibular dysfunction, visual impairment, or other syndromic features were observed. Her parent and sibling were also affected. Although hearing loss was reported across three generations, the presence of multiple consanguineous marriages and three affected individuals within nuclear families suggested an AR inheritance pattern. The proband is married to an unrelated hearing impaired individual (Figure 1A).

### Genomic analysis

Whole-exome sequencing was performed for the proband and their spouse. The proband presented a homozygous splice site variant in NM_001145028.2(*PALM3*):c.314+1G>A and a homozygous variant in NM_144672.4(*OTOA*):c.1939G>C, p.Gly647Arg (Table 1). The *OTOA* variant has been previously published (Sloan-Heggen et al., 2015) and is classified as pathogenic in the Deafness Variation Database and as a variant of uncertain significance (VUS) in ClinVar through manual classification using the hearing loss adjusted ACMG guidelines. The spouse was identified with a homozygous variant in NM_138691.2(*TMC1*):c.1535G>A, p.Arg512Gln. According to the American College of Medical Genetics and Genomics (ACMG) variant interpretation guidelines for genetic hearing loss (Oza et al., 2018), both variants in *OTOA* (PM2_Supporting, PP3_Moderate, PP1_Supporting) and *TMC1* (DFNB7) (PM2_Supporting) were classified as a VUS (Table 1). The *PALM3* c.314+1G>A variant was absent in population frequency databases and predicted to cause abolishment of the native splice donor site unanimously by SpliceSiteFinder-like, MaxEntScan, NNSPLICE, GeneSplicer, SpliceAI, AbSplice, and varSEAK (Figure 1B).

**Table 1.** Variants identified in the present study.

| Gene<br>(Transcript) | c. position | p. position | AF gnomAD<br>(v4.1.0) | MAF gnomAD<br>(v4.1.0) | MAF Pop<br>gnomAD (v4.1.0) | TOPMed<br>v8 | All of<br>Us | SIFT | PP-2 | FATHMM | MT | REVEL | CADD | ACMG Classification |
| --- | --- | --- | --- | --- | --- | --- | --- | --- | --- | --- | --- | --- | --- | --- |
| *PALM3*<br>(NM_001145028.2) | c.314+1G>A | p.? | NP | NP | NP | NP | NP | NS | NS | NS | NS | NS | 35.0<sup>1</sup> | NA |
| *OTOA*<br>(NM_144672.4) | c.1939G>C | p.(Gly647Arg) | 3.01e-6 | 1.7e-4 | Middle Eastern | NP | NP | T | PrD | D | D | D | 24.7<sup>2</sup> | VUS (PM2_Supporting,<br>PP3_Moderate) |
| *TMC1*<br>(NM_138691.2) | c.1535G>A | p.(Arg512Gln) | 5.0e-6 | 3.3e-5 | Admixed American | 4.0e-6 | 1.4e-5 | T | PrD | D | D | U | 33.0<sup>1</sup> | VUS (PM2_Supporting) |

Abbreviations: AF, allele frequency; B, benign; chr, chromosome; D, Deleterious; F, family; MAF, maximum allele frequency; MT, MutationTaster; NA, not applicable; NP, not present; NS, not scored; Pop, population; PP2, PolyPhen-2; PrD, probably damaging; T, tolerated; U, Uncertain, VUS, Variant of Uncertain Significance. Pathogenicity is represented as <sup>1</sup>deleterious or <sup>2</sup>neutral prediction. PM2, rare in population databases; PP3, computational evidence supports deleterious effect; PP1, co-segregation.

Segregation analysis by Sanger sequencing confirmed that the proband had homozygous *PALM3* and *OTOA* variants. The affected parent was homozygous for the *OTOA* variant and heterozygous for the *PALM3* variant. The unaffected parent is a heterozygous carrier of both *OTOA* and *PALM3* variants. The affected brother was unavailable for testing. The proband’s spouse was confirmed with *PALM3* and *OTOA* reference alleles and hearing loss is suspected to be due to the homozygous *TMC1* variant, despite both variants in known hearing loss-associated genes are classified as VUS (Figure 1A).

### Functional studies

A minigene assay evaluated the effect of the *PALM3* c.314+1G>A variant on splicing. RT-PCR demonstrated aberrant splicing consistent with exon skipping (Figure1 C, D). Skipping of exon 4 results in a frameshift (r.173_315del, causing a frameshift (p.Arg58ThrfsTer47), while skipping of exons 4-5 would cause an in-frame deletion (r.172_399del, p.Arg58_Gln133del). All known *PALM3* transcripts are predicted to be affected (Figure 1E).

Consistent with these findings, a previously published *Palm3*-KO mouse exhibits auditory dysfunction with elevated ABR thresholds (Halim et al., 2025). To assess whether reduced gene dosage might contribute to age-related vulnerability, we examined cochleae from 12-month-old heterozygous mice. Confocal imaging of the organ of Corti revealed preserved overall architecture in heterozygous mice compared with wild-type controls (Figure 2A–B). Quantification of IHCs and OHCs showed comparable cell counts between genotypes at (Figure 2C), suggesting that partial loss of *Palm3* does not increase age-related hair cell susceptibility. Together, these results support a LOF mechanism for *PALM3*-associated hearing loss.

**Figure 2.**
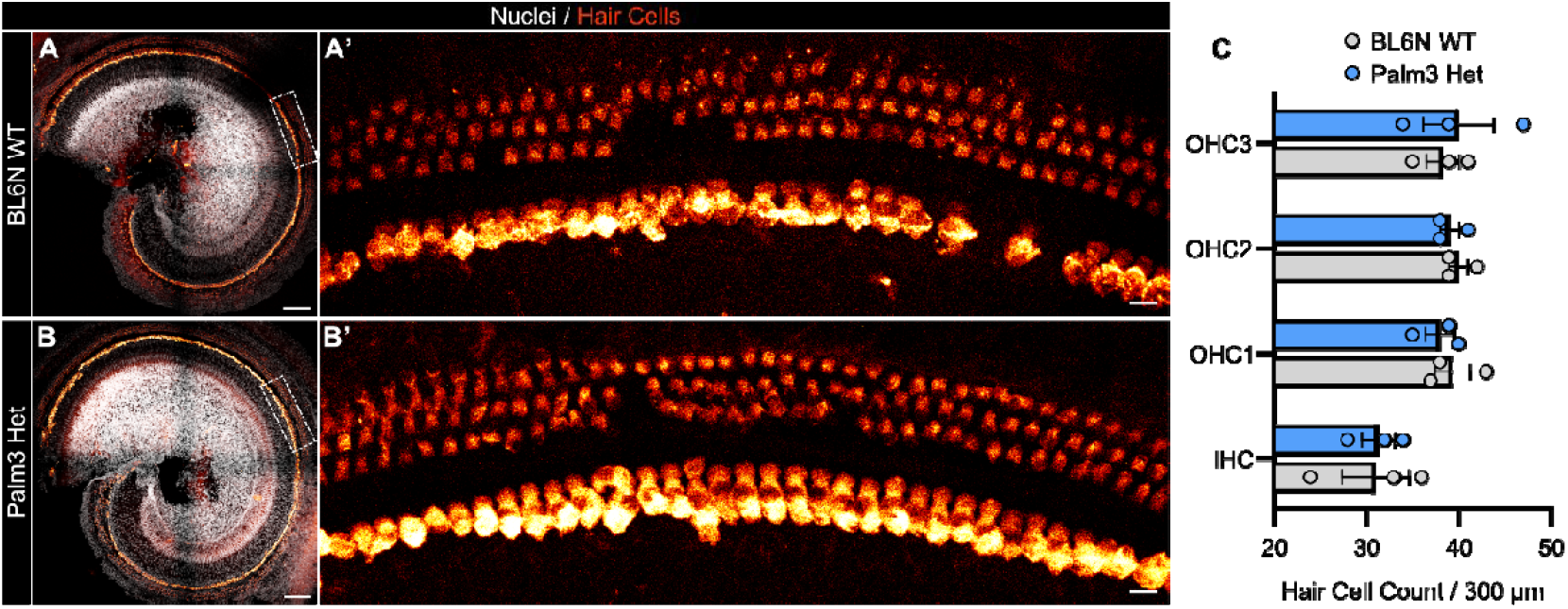
Outer and inner hair cell counts and morphology in the aged *Palm3* heterozygous mouse. (A, A’, B, B’) Representative maximum projections of confocal z-stacks from organs of Corti of 1-year-old BL6N wild-type (A) and *Palm3* heterozygous (B) mice, with indicated hair cell regions magnified in A’ and B’. Tissue was stained for hair cells and nuclei using Myosin7A (red) and TO-PRO-3 (gray), respectively. Scale bars: 100 µm (A, B) and 10 µm (A’, B’). (C) Bar graph showing mean ± SEM hair cell counts of inner hair cells and first, second, and third rows of outer hair cells (IHC, OHC1, OHC2, OHC3) in BL6N WT and *Palm3* heterozygous animals. N = 2 animals per genotype; n = 3 organs of Corti analyzed in total.

## DISCUSSION

Many hearing loss-associated genes that remain to be discovered are expected to be exceptionally rare and are most effectively identified through studying underrepresented populations and consanguineous families, which may harbour multiple homozygous loci (Owrang et al., 2025). While we initially suspected the proband to be diagnosed due to the homozygous *OTOA* c.1939G>C, p.(Gly647Arg) variant, reanalysis uncovered the homozygous *PALM3* c.314+1G>A variant that demonstrated aberrant splicing with two potential splice products. Exon 4 skipping would cause a frameshift (r.173_315del, (p.Arg58ThrfsTer47), while skipping of exons 4-5 would cause an in-frame deletion (r.172_399del, p.Arg58_Gln133del) removing 75 amino acid residues in the longest isoform encoding 688 amino acids (NM_001145028.2). Both outcomes are consistent with loss of normal protein function.

Although *PALM3* does not yet have an established gene-disease relationship in humans, the available genetic and functional evidence suggests that this variant contributes to the proband’s phenotype alongside the segregating *OTOA* variant, representing a potential dual molecular diagnosis in the proband involving two genes that would present as isolated hearing loss. Reporting such cases is important, particularly given the recently characterized mouse model and rarity of families presenting biallelic *PALM3* variants. The confirmed isolated hearing loss in the proband supports that biallelic *PALM3* disruption causes non-syndromic hearing loss and that this gene is likely to gain DFNB designation following the identification of further families segregating biallelic *PALM3* variants.

Genetic factors are estimated to account for approximately 50-70% of variability in auditory acuity in mid⍰ to late⍰life, underscoring the role of inherited susceptibility in ARHL (O’Leary et al., 2025). Increasing evidence suggests overlap between genes implicated in congenital or early-onset deafness and those contributing to ARHL susceptibility (Wells et al., 2019; Vona et al., 2021). Recent large-scale human genetic analyses identified *PALM3* as associated with ARHL (Cornejo-Sanchez et al., 2025), suggesting that rare functional variants may influence auditory decline across the lifespan. Three stages of hearing phenotypes were defined, including self-reported hearing difficulty, hearing difficulty in background noise, and hearing aid use. Rare variants were filtered based on minor allele frequency and predicted functional impact, such as LOF and missense changes. A burden test and Sequence Kernel Association Test – Optimal (SKAT-O) were performed to evaluate the combined impact of variants within each gene. These variant analyses identified *PALM3* associated with hearing aid use (Cornejo-Sanchez et al., 2025).

Paralemmins are membrane-associated proteins involved in membrane dynamics and cytoskeletal interactions (Kutzleb et al., 1998). *PALM3* is expressed in both IHCs and OHCs (Ciuman, 2009). Disruption of membrane and cytoskeletal stability is a mechanism of hair cell vulnerability (Moser et al., 2019). Although ABRs were not performed on the heterozygote mice, and aging to 12 months may not be old enough to model ARHL in mice, our observations, including a biallelic LOF variant in a human affected with ARHL, aberrant splicing demonstrated *in vitro*, and auditory dysfunction in the *Palm3*-KO mouse, support a LOF effect. A recent study, highlighted reduced gene expression as sufficient to influence disease risk in genes that are sensitive to dosage changes (Rice and McLysaght, 2017).

While the contribution of the *PALM3* variant remains masked by the *OTOA* variant in the proband, the combined segregation, functional, and comparative model data strengthen its candidacy. These findings emphasize *PALM3* as a promising contributor to hereditary hearing loss, target for further functional and therapeutic studies and underscore the importance of continued analysis in families with VUS.

## Data Availability

All data produced are available online at ClinVar under accession SUB16130109. Original data produced in the present study are available upon reasonable request to the authors.

https://www.ncbi.nlm.nih.gov/clinvar/

## Acknowledgments

The authors express their gratitude to the family for their kind participation in this study. This work was supported by the Center for Rare Hearing Disorders at the Center of Rare Diseases Göttingen (ZSEG). This work was supported by the German Research Foundation (DFG) VO 2138/7-1 grant 469177153, the DFG Heisenberg program VO 2138/8-1 grant 543719215, and the DFG Collaborative Research Center 1690 (Project A03) to BV as well as the Austrian Science Fund (10.55776/PIN5995324) to CV.

## Author contributions

Conceptualization: T.H., R.M., B.V.; Data curation: P.N.T., L.H., C.V, B.V.; Formal analysis: L.H., R.M., C.V., B.V. ; Investigation: P.N.T., L.H., M.A.H.H, R.M., C.V., B.V.; Methodology: E.G.K, R.M., T.H., C.V.; Resources: M.W.K, E.G.K, R.M., T.H, C.V., B.V.; Writing-original draft: PNT, C.V, B.V.; Writing-review & editing: P.N.T., L.H., M.A.H.H., M.W.K., E.G.K., R.M., T.H., C.V., B.V.

## Declarations

### Ethical approval

The ethics committee of the Medical Faculty of the University of Würzburg gave ethical approval for this work (approval numbers 131/21, 226/18 and 46/15).

### Data availability

The *OTOA* and *TMC1* variants have been submitted to ClinVar under accession SUB16130109.

