## Supplementary Table 1 for "*PALM3* and hearing loss: a potential dual diagnosis interfering with novel gene discovery"

**Supplementary Table 1.** Primers used for segregation testing and *in vitro* splice assay

| Name | Primer sequence (5’-3’) | Amplicon size (bp) |
| --- | --- | --- |
| hu PALM3 Ex4-5 XhoI F | aattctcgagTCTGGGACAGTGATGCTGTT | 864 bp |
| hu PALM3 Ex4-5 BamHI R | attggatccTGACTTGCCAAGACCACAGA |  |
| OTOA Ex19 F | GAATGTCTTTGGGATGAGCTCTTGG | 378 bp |
| OTOA Ex19 R | TTTAGAGTAAGCTTCAAGACCCACC |  |
| TMC1 Ex 16-17 F | CCTAGTTTCTCCCTTGTGCCT | 643 bp |
| TMC1 Ex16-17 R | AATTCAGAGCCAGCACACAG |  |
| SD6 F | TCTGAGTCACCTGGACAACC | NA |
| SA2 R | ATCTCAGTGGTATTTGTGAGC |  |
